# Artificial intelligence for aortic valve calcium score quantification by echocardiography

**DOI:** 10.64898/2025.12.26.25343075

**Authors:** Pere Lopez-Gutierrez, Alberto Morales-Galán, Laura Galian-Gay, Juan Garrido-Oliver, Lydia Dux-Santoy, Neil J Craig, Zi Ye, Josep M. Alegret, Javier Bermejo, Francisco Calvo-Iglesias, Elena Ferrer-Sistach, Irene Méndez, Juan M. Robledo Carmona, Violeta Sanchez, Daniel Saura, Teresa Sevilla, Thomas A. Foley, Hug Cuellar-Calabria, Ignacio Ferreira-González, Hector I. Michelena, Marc R Dweck, Arturo Evangelista, Gisela Teixido-Tura, Jose F. Rodríguez-Palomares, Andrea Guala

**Author notes:** **Corresponding authors:** Andrea Guala. Vall d’Hebron Institut de Recerca, Passeig de la Vall d’Hebron, 129, 08035 Barcelona, Barcelona, Spain.

## Abstract

**Background:** Aortic valve calcification (AVC), as measured by gold-standard computed tomography (CT) Agatston score, provides an anatomic assessment of aortic stenosis (AS) severity and is a key predictor of AS progression and need for valve replacement. AVC detection and quantification from transthoracic echocardiography (TTE) could expand AS early diagnosis and risk stratification, currently limited by CT availability and radiation exposure.

**Methods:** A multi-view video-based deep learning framework was developed using 1166 TTE aortic valve videos from 187 TTE studies acquired in 110 patients with available AVC score by CT. EchoAVC architecture includes a feature extraction model followed by a quality-aware model that aggregates video-level information to obtain patient-level predictions for AVC detection and quantification (score). The framework was validated internally with 173 TTE studies from 86 patients across seven centres, and externally using 430 TTE studies from 280 patients across four different centres. The associations between EchoAVC estimations and AS severity, progression, and need for aortic valve replacement were examined.

**Results:** EchoAVC demonstrated excellent performance in AVC detection (AUROC 0.98, accuracy 94.4%), and quantification (R = 0.64) in the external multi-centre testing set. EchoAVC score was correlated with echocardiographic AS severity descriptors, including aortic valve mean pressure gradient (ρ = 0.749) and peak velocity (ρ = 0.757), and predicted future increase in mean pressure gradient (ρ = 0.382), peak velocity (ρ = 0.433) and calcium score by CT (ρ = 0.650). In 361 patients followed for a median of 3.8 years, 139 underwent aortic valve replacement. Baseline presence and extent of AVC as predicted by EchoAVC showed strong risk-stratification power for aortic valve replacement, remarkably in line with those obtained by CT, and incremental over TTE AS descriptors. EchoAVC was further tested in routine clinical practice images, confirming strong associations with AS severity and progression, including stratification for incident AS in previously unaffected individuals (p<0.001).

**Conclusions:** EchoAVC enables accurate and non-invasive detection and quantification of AVC, offering substantial diagnostic and prognostic value for aortic stenosis progression and need for valve replacement. This technique holds promise as a scalable tool for early detection and clinical management of aortic valve stenosis.

**Clinical Perspective:** *What Is New?:*

* Aortic valve calcification can be detected and quantified on standard, two-dimensional transthoracic echocardiography
* EchoAVC score is associated with echocardiographic metrics of aortic stenosis severity
* EchoAVC predicts progressive increase in aortic stenosis severity and need for aortic valve replacement

*What Are the Clinical Implications?:*

* This openly available deep learning framework can be used to estimate aortic valve calcium in patients at risk of or presenting aortic stenosis to predict aortic stenosis progression
* EchoAVC may help identify patients likely to have high aortic valve calcium and prioritize them for confirmatory CT assessment, thereby supporting earlier identification of calcific aortic valve disease in clinical pathways.

## Introduction

Valvular heart disease represents a growing public health burden [1], [2], [3], with prevalence and severity increasing markedly with age [4]. Among these conditions, aortic stenosis (AS) is the most prevalent, especially in older adults, and is associated with high rates of surgical intervention and mortality [3], [5]. Transthoracic echocardiography (TTE) is essential in the diagnosis, monitoring, and management of AS, serving as a first line tool in routine care [6].

Valvular calcification is a pathological process that drives progressive leaflet stiffening and valve stenosis and serves as a key predictor of AS progression [7], [8] and a potential target for therapy [9]. Quantified by computed tomography (CT) in the form of aortic valve calcium Agatston score (CT-AVC), it is increasingly used to support diagnostic assessment and intervention strategies [10], [11] especially in patients where TTE measurements are discordant. However, the limited availability, radiation exposure and cost of CT scanning limit wide CT-AVC adoption [6], [12], [13], [14].

Recent studies showed that deep-learning models can be effectively trained to reproduce the clinical analysis regularly performed in TTE studies [15], [16], [17]. Furthermore, recent analyses have shown that deep learning models can learn to detect associations previously considered unfeasible by humans, such as the identification of structural heart diseases or regurgitant valvular heart diseases from electrocardiography data [18], [19].

This work presents EchoAVC, a deep learning framework that processes unselected 2D TTE videos to (i) detect the presence of aortic valve calcification (AVC) and to (ii) provide a continuous quantification aortic valve calcium score (EchoAVC score). In a longitudinal study we also investigated whether EchoAVC could serve as a prognostic marker, providing risk-stratification of AS incidence, AS progression and future aortic valve replacement (AVR).

## Methods

The data that support the findings of this study are available from the corresponding author upon reasonable request. Models and weights are freely available at XXXXXX (upon publication).

### Study population and data source

As shown in Figure 1, four cohorts were available for the development and evaluation of EchoAVC, while an additional cohort was retrieved to assess EchoAVC performance in routine clinical practice. To further evaluate consistency across ultrasound systems, a sixth dataset, comprising same-day TTE and point-of-care ultrasound (POCUS) studies, was employed (see supplementary Figure S1).

**Figure 1.**
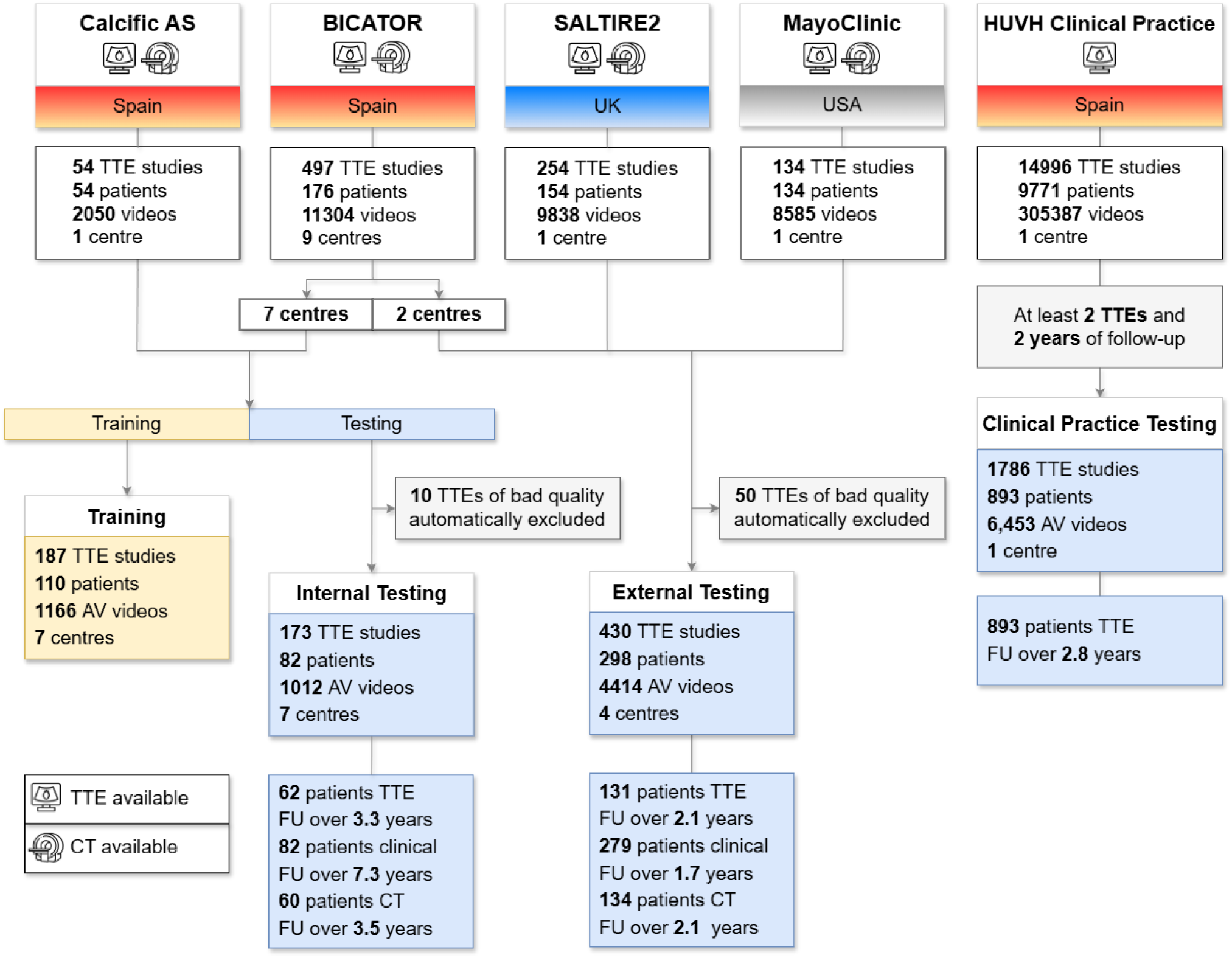
Overview of data available. AS = aortic stenosis, AV = aortic valve, CT = computed tomography, FU = follow-up, HUVH = Hospital Universitari Vall d’Hebron, TTE = transthoracic echocardiography.

### Data for EchoAVC models training

The dataset used for EchoAVC model training comprised two sources of data where all patients underwent both TTE and CT and signed a written, informed consent (Figure 1, left). In both cohorts, CT-AVC was measured on ECG-gated non-contrast CT calcium scoring scans acquired following a standardized protocol and quantified by experienced personnel, and a clinical follow-up was performed in June 2025 to record mortality and incidents of AVR.

The largest dataset was collected for the trial BICATOR, a randomised clinical trial (RCT) that enrolled 220 adult bicuspid aortic valve (BAV) patients across 10 Spanish centres between 2017 and 2021 and that demonstrated that atorvastatin treatment is not effective in reducing AS progression in these patients over 3 years [9]. Key inclusion criteria comprised non-severe valvular dysfunction and calcification. Participants underwent TTE at baseline and annually for 3 years, while CT scans were performed at baseline and after 3 years to quantify CT-AVC. A total of 447 TTE-CT pairs were available. To ensure sufficient representation of tricuspid aortic valves for robust model development, a second dataset, referred to as Calcific AS cohort, was included. It comprises 54 TTE studies, from 54 additional patients with calcific AS enrolled in a previous study at the Hospital Universitari Vall d’Hebron (HUVH) between 2015 and 2020 [20].

After isolating data from 2 BICATOR centres to be reserved for external testing (see below), the remaining dataset was split at the patient level into training and internal testing sets. Thus, 187 TTE studies from 110 patients from 7 Spanish centres were used for model training.

### Data for EchoAVC models testing

From the previously described split of BICATOR and Calcific AS cohorts, 186 TTE studies from 92 patients and 7 centres were available for internal testing of the EchoAVC model.

To assess external validity, model performance was further evaluated on 3 datasets from four clinical centres across three countries, none of which contributed to model training. This external dataset is expected to represent the population attending outpatient cardiac clinics for (risk of) AS, which should thus be considered as the target population of this testing [21]. The first external dataset comprised 89 TTE-CT pairs from 28 patients and 2 centres included in the BICATOR trial. The second external dataset was obtained from the SALTIRE2 RCT [22] and included 254 TTE-CT pairs from 154 participants. Briefly, SALTIRE2 was a single-centre RCT enrolling patients with calcific AS between 2015 and 2017 who were randomised to denosumab, alendronate or placebo, resulting in no difference in the rate of AS progressions between arms. Participants underwent baseline TTE and ECG-gated non-contrast CT, from which CT-AVC was measured following a standardized protocol by experienced personnel. During the 2-year study period, follow-up TTE was performed every 6 months, and non-contrast CT scans were acquired at 12 and 24 months. A clinical follow-up was performed in July 2024 to record all-cause mortality and incident AVR. Written informed consent was obtained from all participants. The third external testing dataset included a total of 134 TTE-CT pairs from 134 patients acquired during clinical care at Mayo Clinic. Among these, 98 patients had calcific AS and BAV, while the remaining 36 presented non-calcific tricuspid aortic valve. All participants received both TTE and non-contrast CT between 2018 and 2025.

In total, 663 TTEs from 408 patients from 7 centres composed the full testing set used for EchoAVC model evaluation, of which 477 TTEs from 316 patients and 4 centres constituted the external test set.

### Data for evaluation in clinical practice and with POCUS

Two additional data sources were included to evaluate the EchoAVC in the context of routine clinical practice and to assess its consistency in POCUS images.

The first, named “HUVH clinical practice cohort”, is a large TTE dataset comprising 14996 TTE studies acquired at HUVH during routine clinical care in 9771 patients between 2019 and 2024. The local Ethics Committee waived informed consent. Among them, 893 patients who received two TTE at least 2 years apart were identified. This cohort was used to (i) test the correlations between EchoAVC score and AS severity metrics as recorded in clinical practice, and (ii) investigate the association between baseline EchoAVC score and the risk of developing AS. For the latter, a subgroup of 99 patients who developed AS (defined as maximum aortic valve velocity > 2.5 m/s) in between the two TTE studies were identified. Then, a propensity score matching (PSM) analysis was performed to identify matched patients that did not develop AS.

The second dataset (named POCUS cohort, Supplementary Figure S1), included 100 consecutive patients referred to HUVH for suspect of cardiac disease and who were enrolled in a research protocol including same-day point-of-care ultrasound (POCUS) and conventional TTE at HUVH in 2025. All patients signed informed consent. This data was used to test the generalizability of the EchoAVC framework to point-of-care ultrasound.

### Data curation and pre-processing

A complete description of pre-processing tasks and models development is included in the supplementary material (see Ancillary models development). Briefly, pre-processing tasks included the automatic identification of TTE views including the aortic valve, an automatic assessment of image quality, the detection of the aortic valve and the subsequent image cropping to specifically focus on the aortic valve. While a pretrained TTE view classifier [23] was used to detect parasternal long-axis, parasternal short-axis, and apical 3-chamber views, two deep learning models, one for image quality and another for valve detection, were trained and tested using a randomly-selected subset of the HUVH clinical practice cohort, comprising 496 TTE studies from 488 patients (Figure S1).

Using these models, low-quality studies were excluded for evaluation of the EchoAVC model through a two-step automatic quality control. First, using the automatic image quality model, each study quality was classified as good, normal or bad quality using the highest video quality within the study. Second, a quality assessment was performed using the image view identification probability, where studies with a mean probability below 80% were classified as bad quality. Studies identified as bad quality in both checks were automatically excluded, with no human intervention.

### EchoAVC model

Inspired by previous artificial intelligence (AI) models in digital pathology and TTE [24], [25], EchoAVC model was developed following a two-stage modelling framework. It includes a first stage that represents the input video in a compact, vectorial form (an encoding, here referred to as “feature extraction model”) and a second stage that aggregates the encoding from multiple videos to obtain patient-level predictions. Models training was performed on a high-performance computing cluster using a set of NVIDIA H100 graphics processing unit (GPU) with mixed precision for training. Full implementation details are included in the supplementary material.

### Feature extraction model

The PanEcho model, a multi-task, video-based deep learning model comprising an image encoder, a temporal transformer, and task-specific output heads [15], was adapted and fine-tuned. In particular, the original prediction heads were removed, and the encoder was fine-tuned by further training of the last two layers after adding several, task-specific heads related to AVC detection and quantification. These tasks included both two- and multi-class AVC classification as well as a regression. AVC prediction heads were removed after the encoder was fine-tuned. The final hidden layer consisted of a 768-element vector, serving as a video-level representation to be used as input for the next model stage. A random five-fold cross-validation approach was used for training and the best-performing model was chosen to extract video-level features. Each fold was trained using input sequences of 32 consecutive frames with a resolution of 224×224 pixels, a batch size of 16, and an SGD optimizer with an initial learning rate of 1e-5.

### Aggregator model

The aggregator model architecture comprised a transformer and two fully connected layers to detect and quantify AVC. To train this model, 30 video-level vectors were combined into a matrix. Each video-level vector included the 768 features extracted from the first stage, along with six additional metadata features: a unique within-study video identifier, the three outputs of the image quality model, and two view-related features (the view and its probability as predicted by the view identification model). For each TTE study, a variable number of matrices were generated depending on the number of videos and frames included in the study to capture information across all aortic valve videos. Predictions for AVC presence and quantification were then aggregated by averaging.

Training and validation were performed with the same data used in the best-performing fold of the first stage. Within this setup, the model achieving the highest testing performance was selected. Training was performed using a batch size of 64 and the AdamW optimizer with a learning rate of 1e-5. Full implementation details are included in the supplementary material.

### Model interpretation

The focus of the model was assessed using saliency maps generated through the Gradient-weighted Class Activation Mapping (Grad-CAM) technique [26]. This method provides visual explanations by highlighting the regions in each video frame that most influenced the model predictions. Specifically, Grad-CAM was applied to the last convolutional block of the EchoAVC feature extraction model, producing heatmaps that were interpolated to the original input resolution.

### Statistical analysis

Categorical variables are reported as counts (valid percentages), and continuous variables as medians (interquartile ranges (IQR)). The AVC detection task was evaluated using accuracy and AUROC, which is equivalent to the c-statistic [21]. Ninety-five percent confidence intervals were calculated using 10000 bootstraps (samples with replacement of the same size as the evaluation set). Brier score and Cox-Snell R2 were computed to evaluate model calibration and overall predictive performance [21], while calibration plots were generated to visually assess the agreement between predicted probabilities and observed outcomes. EchoAVC score predictions were evaluated using the median and IQR of the errors, and the Pearson correlation coefficient (R), and a Bland-Altman plots were generated.

Pairwise correlations between EchoAVC score or ordinal variables clinical variables were assessed using the Spearman rank correlation coefficient. For error sensitivity analyses, absolute errors were compared between two groups using the Mann-Whitney U test. To evaluate residual differences between matched groups in the AS incidence analysis as well as differences in EchoAVC predictions, Mann-Whitney U tests were applied to continuous variables. Time-to-event analyses for incident AVR were performed using a multivariable Fine-Gray proportional subdistribution hazards regression model, considering death as a competing risk. For the incident AS analysis, propensity scores were estimated using logistic regression based on age, time interval between TTE studies, and baseline aortic valve peak velocity. Matching was conducted without replacement, selecting the most similar patient pairs while forcing exact matches for sex and valve morphology (bicuspid vs tricuspid aortic valve). All analyses were performed using Python (version 3.11.9) and R (version 4.5.0) and a p-value ≤0.050 tests was considered significant.

## Results

### Data Overview and Availability

Demographic and clinical characteristics of the patients included are presented in Table 1. The cohort available for model training consisted of a total of 1166 aortic valve videos from 187 TTE studies acquired in 110 patients (Figure 1). Thirty-two patients (29.3%) were women, the median age was 53 years, and the interquartile range was (44-72) years, and 65 patients (59.1%) had a BAV. AVC was absent at both baseline and final TTE scans in 37 patients (33%). Among those with detectable calcification by CT, the median CT-AVC was 1371 (687-3112) AU. The median interval between TTE and CT was 40 (7-134) days.

**Table 1.** Demographic and clinical characteristics of the patients. AV: aortic valve; AS: aortic stenosis; CT-AVC: computed tomography aortic valve calcium score; BAV: bicuspid aortic valve; FU: follow-up; IQR: Interquartile range; HUVH: Hospital Universitario Vall d’Hebron; Vmax: Peak velocity.

|  | Training | Internal Testing | External Testing | HUVH Clinical Practice Testing |
| --- | --- | --- | --- | --- |
| **Study Period** | 2015-2021 | 2015-2021 | 2015-2025 | 2019-2021 |
| **Centres, N** | 7 | 7 | 4 | 1 |
| **Unique patients, N** | 110 | 92 | 316 | 893 |
| **Unique studies, N** | 187 | 183 | 480 | 1786 |
| **Unique videos, N** | 5507 | 5247 | 21023 | 36434 |
| **Age, median (IQR), y** | 55 [43-76] | 42 [37-54] | 67 [27-74] | 67 [52-75] |
| **Sex, N Male (%)** | 78 (71%) | 68 (74%) | 213 (67%) | 545 (61%) |
| **BAV, N (%)** | 65 (59%) | 83 (92%) | 141 (47%) | 99 (11%) |
| **Aortic Stenosis (AS), N (%)** |  |  |  |  |
| None (AV Vmax <=2.5m/s) | 62 (56%) | 71 (77%) | 66 (21%) | 698 (78%) |
| Mild (AV Vmax 2.6-2.9m/s) | 4 (4%) | 4 (4%) | 45 (14%) | 86 (10%) |
| Moderate (AV Vmax 3.0-4.0m/s) | 7 (6%) | 6 (7%) | 160 (51%) | 92 (10%) |
| Severe (AV Vmax >=4.0m/s) | 30 (34%) | 11 (12%) | 45 (14%) | 17 (2%) |
| **AV Area, median (IQR), cm2** | N/A | N/A | 1.0 [0.9-1.2] | N/A |
| **AV Pressure gradient, median (IQR), mm Hg** | 12 [6-45] | 7 [5-12] | 27 [18-33] | 15 [9-23] |
| **AV Vmax, median (IQR), m/s** | 2.1 [1.64-4.11] | 1.74 [1.40-2.36] | 3.40 [2.90-3.80] | 1.54 [1.20-2.36] |
| **AVC Presence (CT-AVC>0), No (%)** | 75 (68%) | 44 (48%) | 265 (84%) | N/A |
| **CT-AVC, median (IQR), AU** | 2708 [1520-4172] | 653 [255-2133] | 1368 [656-2240] | N/A |
| **Patients Echocardiography FU, N (%)** | N/A | 63 (68%) | 178 (56%) | 893 (100%) |
| **Duration, median (IQR), y** | N/A | 3.1 [3.0-3.4] | 2.2 [2.1-2.2] | 2.8 [2.3-3.2] |
| **Delta AV Vmax, median (IQR), m/s** | N/A | 0.03 [0.00-0.22] | 0.41 [0.13-0.64] | 0.07 [-0.13-0.28] |
| **Delta AV Pressure gradient, median (IQR), mm Hg** | N/A | 0.0 [0.0-2.0] | 4.3 [1.3-8.0] | 3.0 [0.0-9.0] |
| **Patients Computed Tomography FU, N (%)** | N/A | 65 (74%) | 134 (42%) | 0 (0%) |
| **Duration, median (IQR), y** | N/A | 3.1 [3.0-3.3] | 2.1 [2.1-2.2] | N/A |
| **Delta CT-AVC, median (IQR), AU** | N/A | 0 [0-189] | 358 [160-832] | N/A |
| **Patients Clinical FU, N (%)** | N/A | 88 (96%) | 316 (100%) | 0 (0%) |
| **Duration, median (IQR), y** | N/A | 7.7 [4.9-8.1] | 7.2 [1.7-8.0] | N/A |
| **AVR, N (%)** | N/A | 20 (22%) | 165 (52%) | N/A |
| **Death, N (%)** | N/A | 8 (9%) | 66 (21%) | N/A |

After the automatic exclusion of 60 studies of low quality (9%), the full testing cohort included 5426 aortic valve videos from 603 TTE studies acquired in 380 patients, comprising 298 patients from the external cohorts. Overall, 119 patients (31%) were women, with a median age of 69 (54-78) years, and 217 patients (57%) had a BAV. AVC was present in 285 patients (75%), with a median CT-AVC of 1128 (520-2155) AU. The median interval between TTE and CT in this cohort was 0 days (0-32).

### EchoAVC evaluation

In the internal testing cohort EchoAVC detection demonstrated excellent discrimination in differentiating the presence or absence of AVC, achieving an AUROC of 0.94 (95% CI: 0.90–0.97) (Supplementary Figure S3A). In the external testing cohort, including 430 studies from 4 centres, EchoAVC detection maintained high performance, with an AUROC of 0.98 (95% CI: 0.96–0.99), an accuracy of 94.4% (Figure 2A) and sensitivity, specificity, and negative and positive predictive value were 0.96, 0.87, 0.85, 0.97. In terms of calibration, Brier scores was 0.05, and calibration plot (Figure 2B) further confirmed the accuracy of calibration. Furthermore, Cox-Snell R2 was 0.45, indicating strong explanatory power.

**Figure 2.**
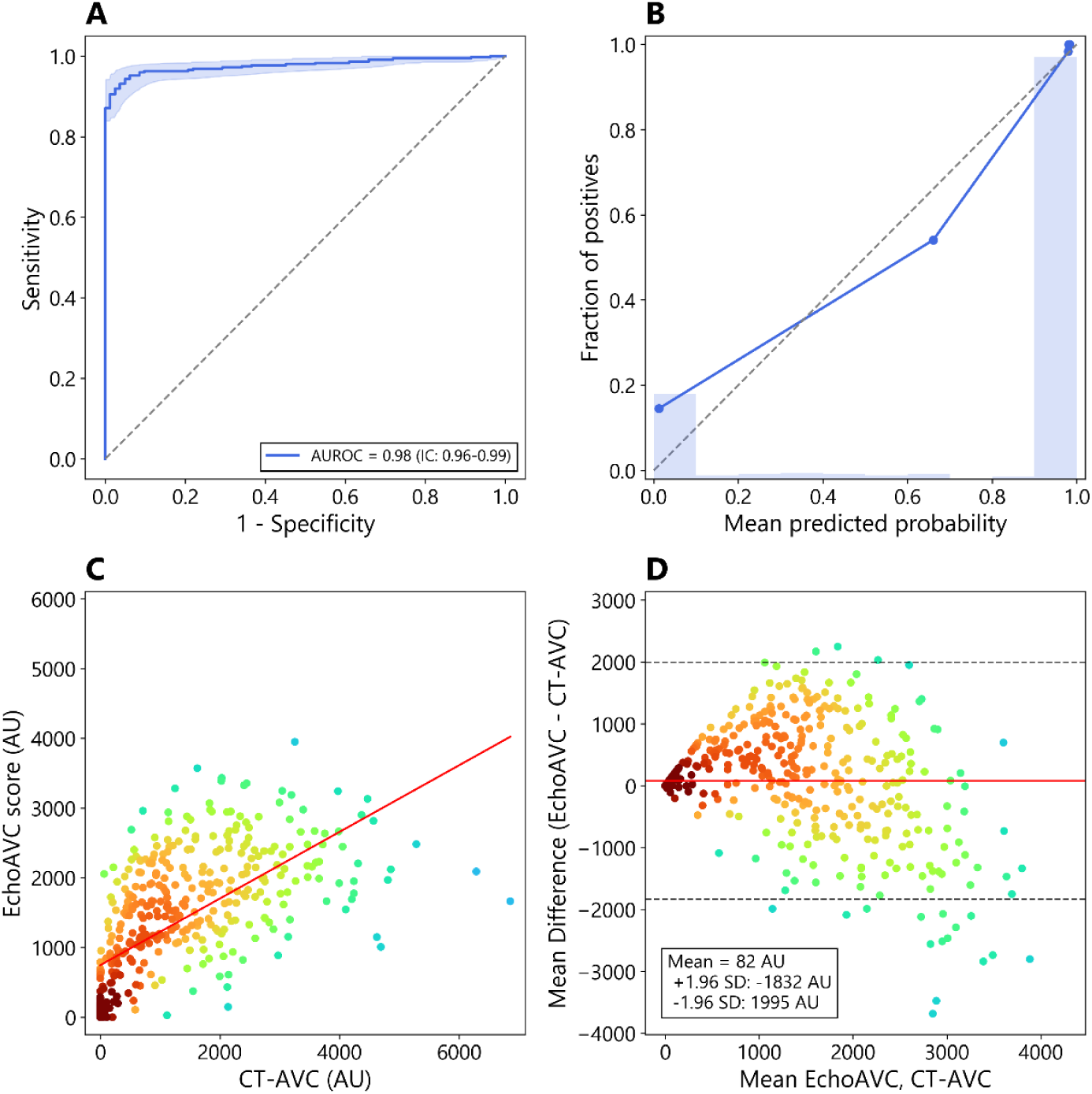
Receiver operating characteristic (ROC) curve (A) and calibration plot (B) for the EchoAVC presence predictions and scatter (C) and Bland-Altmann plot (D) of EchoAVC score in the external testing set. Color represents local point density.

Strong agreement was observed between EchoAVC score and CT-based Agatston calcium score in the multi-centre internal testing set, with a Pearson correlation coefficient of R = 0.78 and a median error of 0 AU (-107 – 62) (Supplementary Figure S3B). In the external testing dataset, the model generalised well, yielding R = 0.64 with a median error of 129 AU (-257 - 649) (Figure 2C) and a minimal bias (82 AU, limits of agreement –1832 to 1995 AU) (Figure 2D). A sensitivity analysis showed no differences in prediction errors comparing female and male patients (Supplementary Figure S4A). Conversely, prediction errors were inversely related to image quality (Supplementary Figure S4B).

### Evaluation of associations with AS severity and progression

In a subset of the testing cohort with available TTE markers of AS, higher predicted EchoAVC scores were cross-sectionally associated with worse indicators of AS severity (Figure 4). Specifically, EchoAVC score was positively correlated with aortic valve peak velocity (ρ = 0.757, p < 0.001) and mean pressure gradient (ρ = 0.749, p < 0.001), and negatively associated with aortic valve area (AVA) (ρ = –0.282, p < 0.001) (Figure 4A, C; Supplementary Figure S5A; Supplementary Table S1). Notably, very similar associations were observed for CT-AVC (Figure 4B, D; Supplementary Figure S5B).

**Figure 4.**
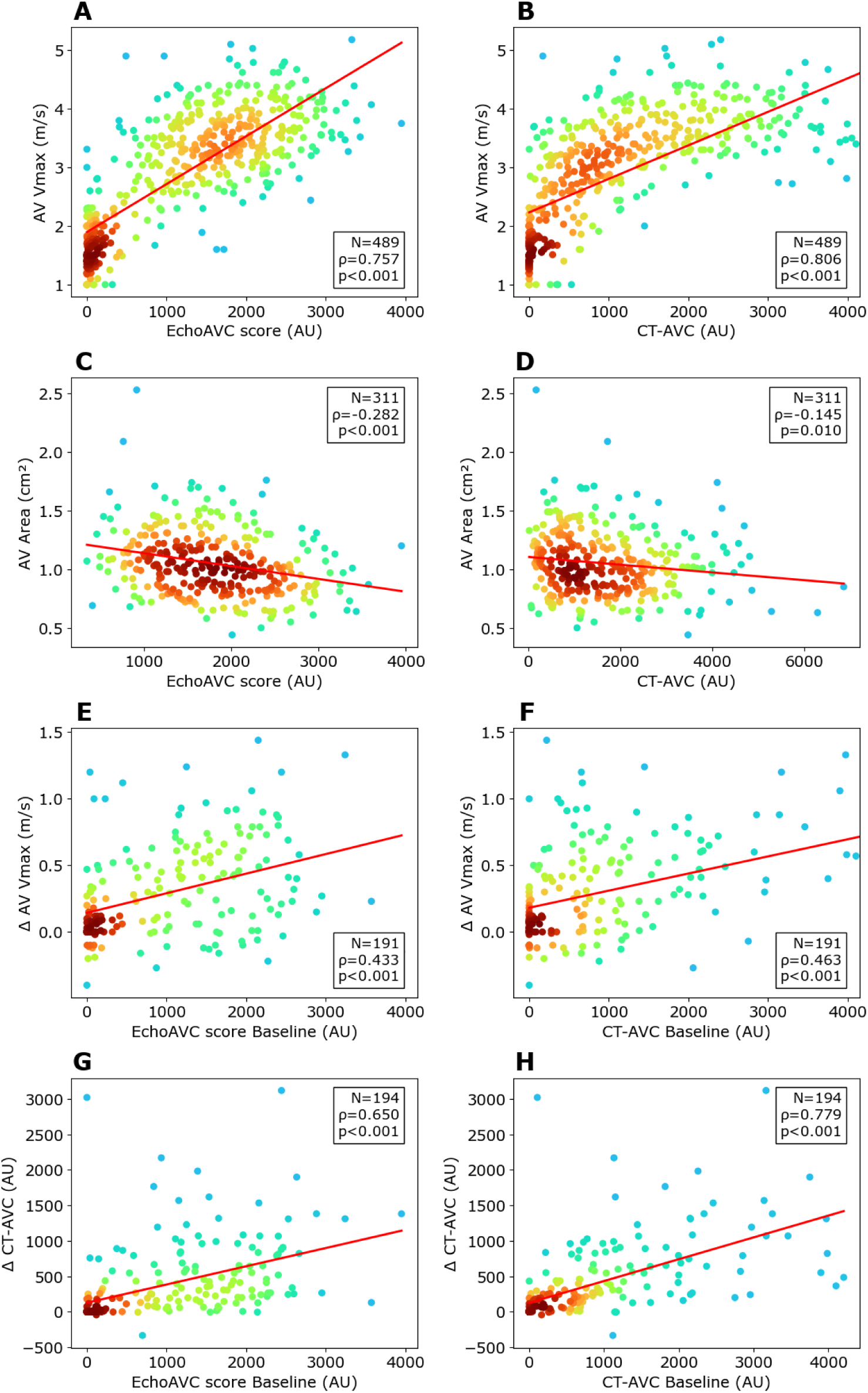
Cross-sectional (A-D) and longitudinal (E-H) associations of aortic valve calcification by EchoAVC (left) and CT-AVC (right) with AS markers. Δ refers to change during follow-up (final – baseline values)

Furthermore, in a separate subset of the testing cohort with longitudinal TTE or CT follow-up (median follow-up of 2.2 [2.1 - 3.1] years), higher baseline EchoAVC scores were positively correlated with future increases in aortic valve peak velocity (ρ = 0.433, p < 0.001), mean pressure gradient (ρ = 0.382, p < 0.001), and CT-AVC (ρ = 0.650, p < 0.001) (Figure 4E, G; Supplementary Figure S5C). Again, these associations were very similar to those obtained with baseline CT-AVC (Figure 4F, H; Supplementary Figure S5D). All Spearman correlation coefficients are provided in Supplementary Table S1. Of note, the associations between baseline EchoAVC and future increases in aortic valve peak velocity and mean pressure gradient were significant even after correcting for baseline aortic valve peak velocity or mean pressure gradient (p=0.005 and p=0.026, respectively) (Table S2).

### Evaluation of association with future AVR

In a subset of the testing cohort including 361 patients followed for a median of 3.8 (IQR 0.4 – 7.1) years, 139 (39%) patients underwent AVR, and 53 (15%) died. EchoAVC presence prediction demonstrated powerful stratification for future AVR (hazard ratio (HR) of 23.1), showing substantial overlap with CT-based risk stratification (Figure 5A). Furthermore, a clear dose-response relationship was observed when stratifying AVR incidence across terciles of predicted EchoAVC scores (Figure 5B). In particular, patients in middle (T2) and high (T3) EchoAVC score terciles demonstrated significantly higher risk of undergoing AVR compared to those in the lowest tercile (T1), with HR of 16.2 and 29.3, respectively (both p < 0.0001). The risk for incident AVR also increased from T2 to T3 (HR = 1.8, p = 0.0006). These patterns closely mirrored the stratification obtained using CT-AVC, while EchoAVC score remained independently associated with AVR incidence after adjustment for baseline AV peak velocity, mean pressure gradient and presence of bicuspid aortic valve (Table S2).

**Figure 5.**
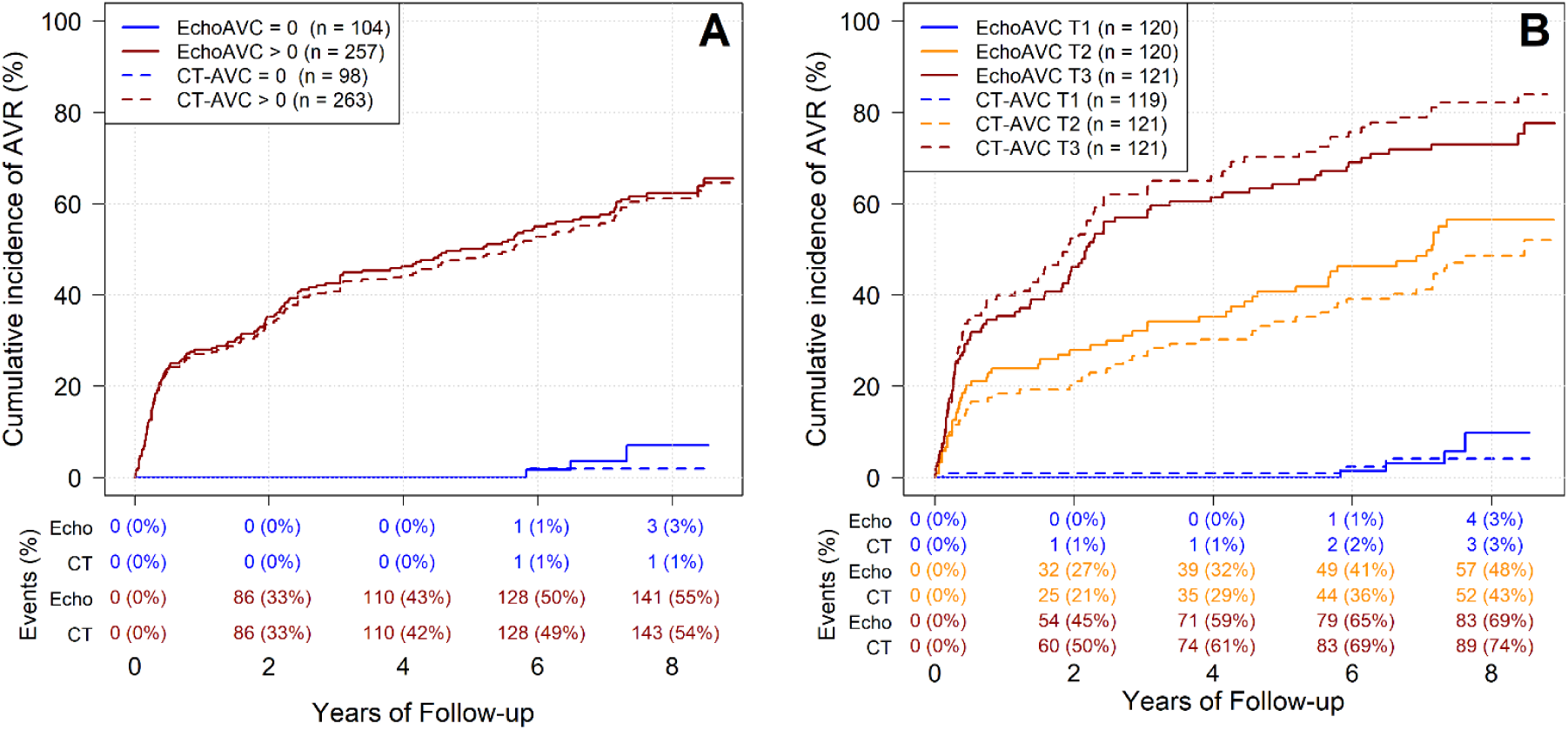
Cumulative incidence of aortic valve replacement (AVR) according to presence or absence of valve calcium (A) or calcium score terciles (B) by EchoAVC and CT-AVC. T = tercile.

### Evaluation of associations in routine clinical practice and consistency in POCUS

To test the generalizability of the associations between EchoAVC score and AS severity and its progression over time in images acquired during routine clinical practice, a total of 893 TTE study pairs acquired a median of 2.8 years apart (IQR: 2.3-3.2) in 893 patients were available from the HUVH clinical practice dataset. Confirming the previously showed results in the testing cohort, EchoAVC score shown positive cross-sectional correlations with aortic valve peak velocity (ρ = 0.547, p < 0.001) and mean pressure gradient (ρ = 0.367, p < 0.001). Furthermore, a higher baseline EchoAVC score was associated with a future increase in aortic valve peak velocity over time (ρ = 0.150, p < 0.001) (Supplementary Figure S6A-C).

In the propensity-score matched clinical practice cohort including a total of 198 patients free from AS at baseline, the 99 patients that developed AS during follow-up presented higher baseline EchoAVC score compared to the 99 patients that did not develop AS (1286 (IQR: 882 – 1778) AU vs 690 (287-1238) AU, p < 0,0001), despite similar baseline age, sex, valve morphology and aortic valve peak velocity (Supplementary Figure S6D and Table S3).

To evaluate the consistency of EchoAVC predictions across ultrasound systems the model was tested on the POCUS cohort, which included 324 aortic valve POCUS videos and 770 TTE videos. EchoAVC detection and score showed strong concordance between POCUS and conventional TTE, with an overall agreement of 83.8% and an AUROC of 0.89 (IC: 0.81-0.95) for classification, and small bias (21 AU) and limits of agreement (–833 - 875 AU) for AVC score predictions (Supplementary Figure S7). This. Of note, POCUS quality was substantially worse than TTE quality (Supplementary Figure S8).

### Prediction interpretability

Grad-CAM was used to identify those regions in each frame that influenced the most the prediction. These saliency maps demonstrated consistent and localized activation in areas corresponding to AVC, whereas patients without AVC showed little or diffuse activation (Figure 6). A video showing grad-CAM at video-level is included in Supplementary Video 1.

**Figure 6.**
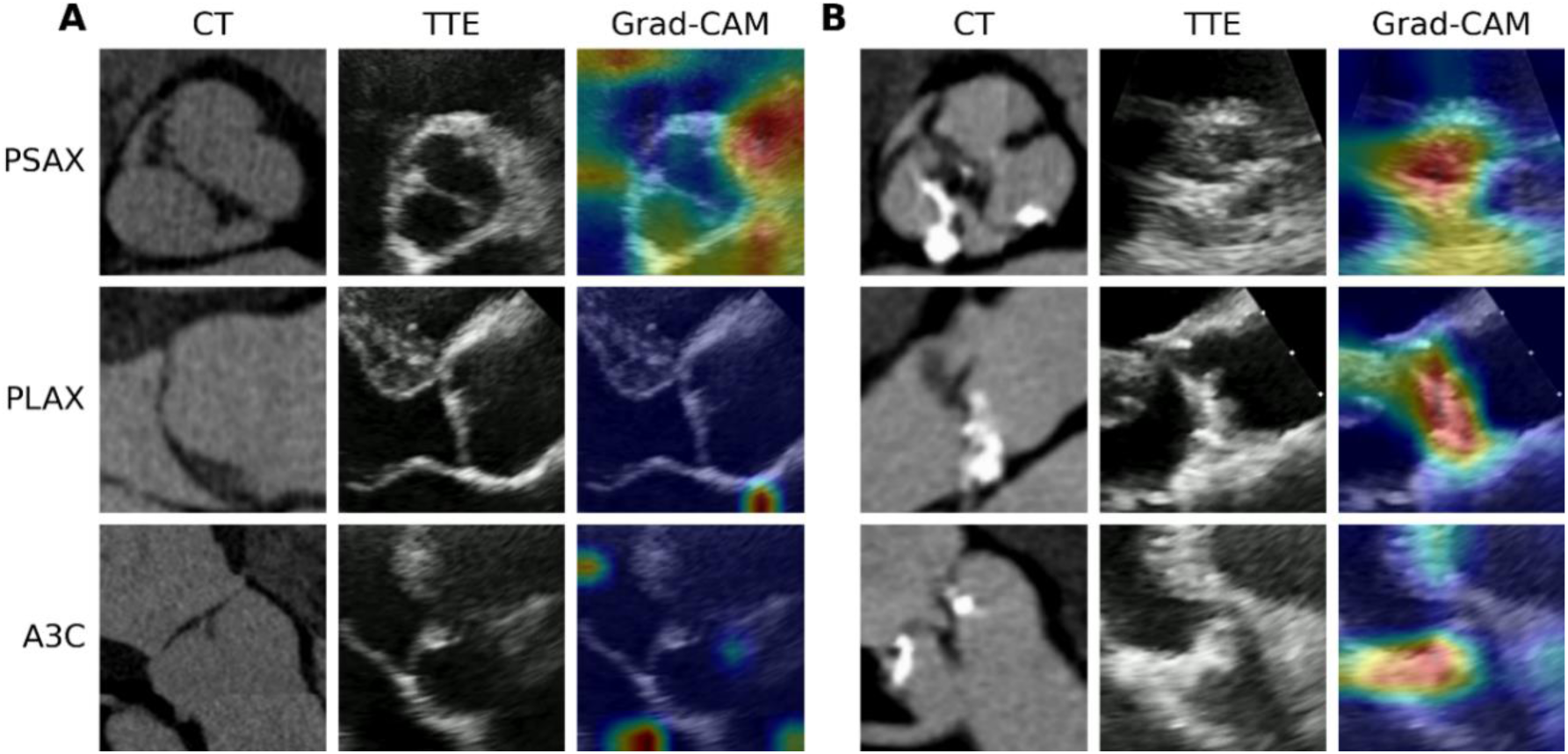
Representative CT, TTE and Grad-CAM images in PSAX (top), PLAX (middle) and apical 3-chamber (bottom) views in a patient with no aortic valve calcium (A) and in a patient with valve calcium (B). Note that while contrast CT are showed, all CT-AVC scores were derived in non-contrast enhanced images.

## Discussion

In this multicentre cohort study, a novel quality-aware deep-learning framework for echocardiography video analysis, EchoAVC, was developed and validated to detect and quantify aortic valve calcification. EchoAVC demonstrated excellent and generalizable performance in detecting AVC presence and quantifying aortic valve Agatston calcium score across diverse populations. EchoAVC predictions further demonstrated relevant diagnostic associations and substantial prognostic implications in predicting incident AS, progressive increase in AS severity, and the need for AVR.

Previous efforts to quantify AVC from TTE presented substantial limitations. Firstly, visual assessment, often based on the visual detection of strongly echogenic spots in the aortic valve, presents limited inter-observer reproducibility [27], [28] and thus is not recommended by clinical guidelines [29]. To improve reproducibility, early semi-automated approaches based on brightness thresholding [30], [31] and more recent deep learning models [32], [33], [34], [35] demonstrated some degree of feasibility in AVC detection in single-centre internal testing sets of very modest size. However, to the best of the authors’ knowledge, all but one study tested their implementations against subjective visual assessment on TTE images. The sole study testing against the gold standard, CT-AVC [31], proposed a thresholding method heavily relying on manual delineation, resulting in relatively low performance in a small (52 patients) internal cohort.

EchoAVC demonstrated excellent performance across diverse populations in detecting the presence of AVC, maintaining AUROC values of 0.98 in the external testing set, confirming its generalizability and transportability. Notably, the model achieved high positive predictive values (0,97) in the external testing cohort, a key performance metric to test its potential utility for clinical screening and early detection of AVC using echocardiography. Beyond AVC detection, EchoAVC provided continuous quantification of AVC score, with a strong correlation to reference CT-based measures. The model presented an amplitude of limits of agreement of 2122 and 3827 AU in the internal and external testing set, respectively. The strengths of these associations should be considered in light of the inherent variability in CT-AVC measurements. To the best of the authors knowledge, only two studies reported scan-rescan inter-observer variability of Agatston calcium score by CT. One study reported limits of agreement amplitude of 855 AU in 15 patients with AS [36], while the other study reported limits of agreement amplitude of 1063 AU in 37 MESA participants with non-zero calcium score [37]. While EchoAVC score errors showed wider dispersion compared to these single-center reports, this may partially reflect the increased inherent complexity of multicentre echocardiographic data as well as of the much larger cohort.

EchoAVC score demonstrated substantial diagnostic and prognostic significance, as predicted baseline score was significantly associated with cross-sectional AS markers and, most importantly, with progression of AS severity and risk of incident AVR. A strong association between EchoAVC score and AS severity was consistently observed across several descriptors of AS severity, including the aortic valve mean pressure gradient, peak aortic valve velocity, and aortic valve area, and in both research and clinical practice datasets. Similarly, the association between baseline EchoAVC score and stenosis progression was demonstrated across internal and external research data as well as in a large clinical practice sample. Notably, these associations closely paralleled those observed with CT-AVC, further underlying the potential of EchoAVC to serve as a non-invasive surrogate for CT-AVC in capturing and tracking AS severity and progression. Finally, the capacity to stratify the risk of incident AVR was notably similar to that obtained by CT-based Agatston calcium score, supporting a potential practical equivalence for predicting this relevant endpoint, while indicating that EchoAVC score provides incremental prognostic value beyond standard echocardiographic parameters.

As expected, prediction errors were significantly higher in images with low-quality compared with images identified as of normal or good quality. This is in line with numerous previous studies relating lower image quality with lower performance and higher inter-observer variability in the measurement of several simple and advanced echocardiography metrics [38], [39], [40], [41]. This underlines the importance of image quality, especially for fine-grain tasks such as AVC quantification and strain analysis and highlights the need for consistent acquisitions [42] and automatic quality control tools.

We anticipate two main possible uses of EchoAVC in clinical practice. First, EchoAVC may play a role in the diagnosis of severe AS in patients with discordant TTE metrics, potentially serving as an alternative to CT in cases where CT is unavailable. Notably, a negative EchoAVC score would almost certainly indicate a negative CT-AVC, meaning that patients could avoid the radiation exposure and cost of a CT scan. This suggests EchoAVC could be integrated into diagnostic pathways to triage patients between routine echocardiographic assessment and CT, particularly in cases of diagnostic uncertainty. In this respect, future studies should define and validate threshold values of EchoAVC that correspond to different stages of AS severity, particularly in challenging cases such as low-flow, low-gradient AS, to enable its full integration as a reliable diagnostic tool for this entity. Secondly, EchoAVC may have utility in population screening, particularly as a scalable, radiation-free tool to detect early signs of valve calcification. In this respect, although the model showed high positive predictive value for AVC detection, it was not directly evaluated in a prospective cohort of asymptomatic patients with relatively low pre-test probability of AS, an important test that should be performed. Additionally, EchoAVC provides a continuous, interpretable score that could be monitored over time, offering a non-invasive method to track structural AS progression or evaluate response to therapy.

## Limitations

Certain study limitations warrant consideration. Firstly, the model was trained and validated exclusively on transthoracic echocardiography data acquired with only two major ultrasound vendors (Philips and General Electric). No data from other commercial systems nor transoesophageal echocardiography was included, leaving generalizability of the framework to other these systems untested. Furthermore, no direct comparison was performed between CT-AVC and predictions on POCUS images. Dedicated testing studies should be performed.

## Conclusions

A novel artificial intelligence implementation accurately detects and quantifies aortic valve Agatston calcium score from echocardiography videos. EchoAVC can prognosticate differences in AS severity progression and future AVR. These findings highlight the potential of EchoAVC as a non-invasive alternative for AVC-based risk stratification.

## Data Availability

The data that support the findings of this study are available from the corresponding author upon reasonable request

## Acknowledgments

We are grateful to José Manuel Crespín Esquivel and Oscar Mula from the IT Department of the Hospital Universitari Vall d’Hebron for their support in the extraction of TTE and to Garazi Urio Garmendia, Eduard Rodenas Alesina, and Augusto Sao Aviles for their fruitful discussions on statistical analysis.

## Non-standard Abbreviations and Acronyms

AS: aortic stenosis
AVC: aortic valve calcium
BAV: bicuspid aortic valve
CT: computed tomography
POCUS: point-of-care ultrasound
TTE: transthoracic echocardiography
Vmax: aortic valve peak velocity

